# Risk and Spatial Spread of a Measles Outbreak in Texas

**DOI:** 10.1101/2025.09.01.25334576

**Authors:** Martial L. Ndeffo-Mbah, Sina Mokhtar, Abhishek Pandey, Chad R. Wells

## Abstract

**Background:** In January 2025, a measles outbreak was reported in Gaines County, Texas, and spread to other counties and states. However, investigations on the risk of the geographical spread of such an outbreak are limited.

**Methods:** In this study, we developed a measles transmission model parameterized with 2020-2024 measles-mumps-rubella (MMR) vaccination coverage and human mobility data in Texas. We compared model predictions to data from the 2025 measles outbreak in West Texas and simulated counterfactual outbreak scenarios in Texas under different vaccine uptake and originating counties.

**Results:** We showed that a measles outbreak originating in Gaines County would have at least an 80% probability of directly generating local outbreaks in six neighboring counties (Terry, Lubbock, Yoakum, Andrews, Ector, and Midland County) in West Texas. When considering second-generation outbreaks, seven additional counties - El Paso, Bexar, Tarrant, Dallas, Harris, Dawson, and Hockley County - had at least a 60% probability of experiencing a local outbreak. Moreover, we showed that an outbreak originating in a low MMR-vaccine county, such as Polk, Montague, and Limestone County, has at least an 80% probability of causing large outbreaks in metropolitan areas such as Houston and Dallas. Moreover, we showed that a statewide increase in vaccine coverage is necessary to prevent large-scale measles outbreaks in Texas.

**Conclusions:** Measles has the potential risk of causing statewide outbreaks in Texas. Our study provides a modeling framework that can be used to inform county-level risk and spread of a measles outbreak and evaluate pre-emptive vaccination strategies.

## Introduction

Measles is an acute, highly contagious respiratory viral disease that typically presents with a blotchy skin rash. The virus infects around 90% of non-immunized individuals exposed to an infectious case [1]. Though measles usually doesn’t cause long-term medical issues, infection can cause severe complications and even death [2]. Following the isolation of the measles virus in 1954, the first measles vaccine was licensed in the United States of America (USA) in 1963[3,4]. Since then, the measles vaccine has undergone refinement, leading to the highly effective and safe measles, mumps, and rubella (MMR) vaccine widely used today [3]. Measles vaccination has dramatically reduced the burden of measles in the United States and globally [5].

In the USA, measles was declared eliminated in 2000, a testament to the vaccine’s success [3]. However, measles outbreaks continue resurfacing in the USA [6]. The country has witnessed sporadic outbreaks in the post-elimination period, often linked to international travel. These outbreaks highlight the importance of sustained vaccination efforts and vigilant surveillance. According to the US Centers for Disease Control and Prevention (CDC), between 2001 and 2019, 158 measles outbreaks were reported in the US, resulting in over 3,000 cases [6]. Most of these outbreaks occurred in communities with low vaccination rates, underscoring the critical role of herd immunity in preventing the spread of measles.

On January 23rd, 2025, the Texas Department of State Health Services (DSHS) reported the first measles cases in the state since 2023 [7]. On January 29th, local transmission among unvaccinated school-aged children was identified in Gaines County, Texas [7]. The outbreak originated in a predominantly Mennonite community with low vaccine uptake. By March 4th, 159 cases were reported in Texas, with 107 cases in Gaines County. Since then, the outbreak has spread throughout West Texas and beyond, with Gaines County serving as the epicenter. By June 6th, 742 cases had been reported in 24 counties out of the 245 counties in Texas, with 55% of cases in Gaines County [8]. In response to this ongoing outbreak, DSHS launched communications, testing, and vaccination campaign efforts to mitigate the spread and burden of measles in Texas.

In this study, we developed a data-driven model to analyze the risk and spatial spread of a measles outbreak across Texas. Our model integrates county-level school immunization data from Texas DSHS and a human mobility dataset derived from anonymized mobile phone records (SafeGraph). Using the 2025 Gaines County measles outbreak as a case study, we estimated the likelihood that an outbreak in this region could spread statewide. We also estimated the potential impact of various vaccination strategies on the risk and spatial spread of the epidemic in Texas.

## Methods

### Transmission risk model

To estimate the risk of a measles outbreak spreading in Texas, we considered the state as a network of counties linked through human mobility. The basis of the model is that the network of contacts between counties can provide estimates of the probability that a county outbreak can seed an outbreak in an interconnected county. For each pair of counties, we calculated the probability that an outbreak could be seeded in county *i* given that an outbreak occurs in neighbouring county *j*. To compute this probability, we first consider the probability of measles transmission through direct contact between an infectious and a susceptible individual as a set value *q*. The probability that an individual in county *j* becomes infected during a local outbreak is denoted by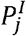, and the probability that an individual in county *i* is susceptible is denoted by 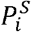 The probability that a single infected individual in county *i* causes a large outbreak in that county is denoted by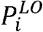. So, the probability that an outbreak in county *j* leads to an outbreak in county *i* through a single contact between the counties is given by 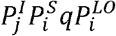.

We assumed 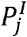 to be equal to the proportion of individuals infected during the outbreak in county *j*. This proportion is considered to be equal to the solution of the final outbreak size equation [9]

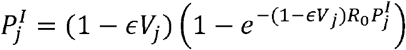

where *R*_*0*_ is the measles basic reproductive number, is the MMR vaccine efficacy, and *V* _*j*_ is vaccine coverage in county *j*. The basic reproductive number is defined as the number of secondary cases caused by a single infective case in a completely susceptible population [10].

We assumed that 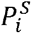 is equal to (1 - *∈ Vi*), which is the proportion of the population unimmunized (unvaccinated or not effectively immunized).

We defined 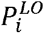 using the Anderson & Watson formula for the probability of a major outbreak of an SEIR-type pathogen in a susceptible population [11,12]

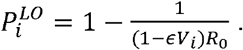

Here, a major outbreak refers to a continuous chain of pathogen transmission with a reproduction number greater than 1 [11,12]. An SEIR-type pathogen is a pathogen, such as the measles virus, whose outbreak dynamics can be modeled using an SEIR (Susceptible-Exposed-Infectious-Recovered) model [13,14].

We set *R*_*0*_ to be equal to 18 [10], and the MMR vaccine efficacy *∈*= 0.97 [15]. We use county-level kindergarten average MMR vaccination coverage from 2020-2024 to inform our model baseline vaccination coverage [16]. From 2020-2024, the average state-level MMR vaccination coverage among kindergartners in Texas was 95%.

If *C*_*ij*_ is the number of contact pairs that link county *i* and *j*, the probability that at least one contact pair causes a major outbreak in county *i* is given

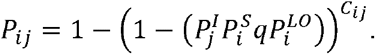

We defined *C*_*ij*_ as the number of visits from county *j* to county *i* during a year. To estimate *C*_*ij*_,we used between-county visitor flows in 2019 computed from anonymous mobile phone users’ visit data provided by SafeGraph [17]. The flows dataset was validated against the American Community Survey commuting dataset [17].

A county outbreak resulting from contacts with infected individuals from the source county (e.g., Gaines County) would be denoted as a first-generation outbreak. Under this definition, a second-generation outbreak is a county outbreak resulting from contacts with infected individuals from a first-generation outbreak county. If we denote by *i* the source county, the expected probability of a second-generation outbreak in county *j* is defined as 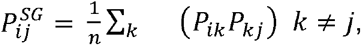, where *k* are potential first-generation outbreak counties, and n is the number of first-generation counties. The expected probability value of a second-generation outbreak, 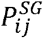, decreases as the number of first-generation counties, *n*, increases. So, accounting for the contribution of low-risk first-generation outbreak counties to this formula may underestimate the expected risk of a second-generation outbreak.

Though the model can be extended to account for higher-order generations such as third- and fourth-generation outbreaks, we limited our model to second-generation outbreaks, given that the bulk of measles cases and active transmission events, in this Texas outbreak, have been limited to a handful of counties: Gaines, Terry, Lubbock, El Paso, and Dawson counties (Table S1).

To minimize the potential ‘dilution effect’ of low-risk first-generation outbreaks on our estimation of the probability of second-generation outbreaks, we limited our formula to counties with a first-generation outbreak probability *P*_*ik*_ greater than or equal to 0.5. Under this assumption, our combined first- and second-generation outbreak probability across West Texas counties was consistent with reported county-level cases, as counties with the highest number of reported cases had a higher predicted outbreak probability (see Results).

### Vaccination scenarios

We consider three vaccination scenarios to estimate the impact of increased vaccine uptake on the risk and spatial spread of a measles outbreak in Texas. These scenarios do not aim at investigating vaccination response strategies to the ongoing measles outbreak, but instead to investigate the potential impact of counterfactual baseline vaccination coverage. These are paramount for informing pre-emptive vaccination strategies for mitigating the risk of future measles outbreaks in Texas. In the first scenario (strategy 1), we assumed that at least 90% of the population is vaccinated in all counties, which is the average MMR vaccination coverage among 24-month-old children in Texas in 2022 [18]. In this scenario, the vaccination coverage of 17 counties is increased to 90%, resulting in a 95.6% state-level vaccination coverage (Fig. S1). In the second scenario (strategy 2), we assumed that at least 92% of the population is vaccinated in all counties. This is the average MMR vaccination coverage among kindergarten children in the USA in 2023-2024 [19]. In this scenario, the vaccination coverage of 22 counties is increased to 92%, resulting in a 95.8% state-level vaccination coverage (Fig. S1). In the third scenario (strategy 3), we assumed that each county’s vaccination coverage would increase by 5% (up to 100% coverage). Here, 94% of Texas counties achieve or exceed the 95% vaccination coverage threshold (Fig. S1), the US CDC Healthy People 2030 MMR target for preventing measles outbreaks [20]. This 5% increase results in a 99% state-level vaccination coverage.

### Alternative outbreak scenarios

We considered alternative scenarios where a hypothetical 2025 measles outbreak in Texas originated in other Texas counties (Fig. S1). We focused on outbreaks starting in counties with low-vaccine coverage, less than 90%, and compared their risk of spatial spread throughout the state to the ongoing outbreak that originated in Gaines County. We specifically focused on King County, Hall County, and Throckmorton County, which were identified as the lowest vaccine coverage counties in Texas, with 80.6%, 82.0%, and 83.4% coverage, respectively. These three counties are sparsely populated, low-population-density rural counties, isolated from large Texas metropolitan areas. In addition, we considered low vaccine coverage counties such as Limestone County, Polk County, and Montague County, with 86.4%, 86.8%, and 89.2% coverage, respectively. These counties are situated in proximity to the largest metropolitan areas, which are Houston and Dallas-Fort Worth.

## Results

Our simulations show that a 2025 measles outbreak originating in Gaines County would likely spread to neighbouring counties and generate intra-county outbreaks (large-scale local transmission) mostly in West Texas (Fig. 1). West Texas counties such as Lubbock, Terry, Yoakum, Andrews, Ector, and Midland had a high probability (above 0.8) of experiencing a local outbreak directly initiated from the Gaines County outbreak, whereas Dawson County had a moderate risk, around 0.5 (Fig. 1A).

**Figure 1.**
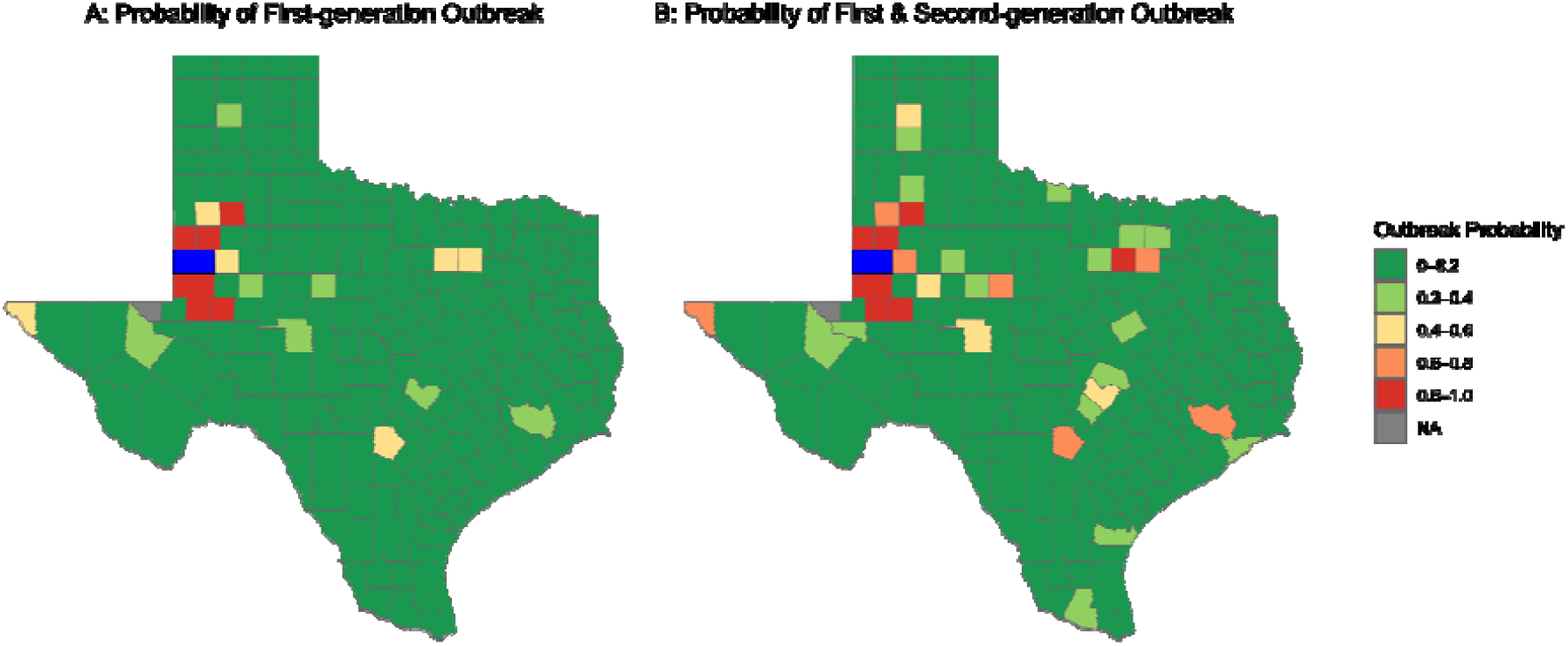
Probability that a measles outbreak originating in Gaines County leads to major outbreaks in other Texas counties. Gaines County, the source of the outbreak, is denoted in blue. A: Probability of first-generation outbreak: i.e., outbreaks directly seeded by infectious individuals traveling from Gaines County. B: The probability of first- and second-generation outbreaks; i.e., outbreaks initiated either directly from Gaines County or first-generation affected counties. The county shapefile was obtained from the US Census Bureau

When accounting for both first- and second-generation outbreaks, 13 (Lubbock, Terry, Yoakum, Andrews, Ector, Midland, Dawson, Dallas, Tarrant, Bexar, El Paso, Harris, and Hockley County) counties showed at least a 0.6 probability of experiencing a local outbreak (Fig. 1B). We found that densely populated counties such as Dallas and Tarrant County in North Texas, Bexar in South Texas, and El Paso in West Texas faced a 0.6-0.8 probability of experiencing a local outbreak through direct contact with Gaines County or contacts with Counties directly infected by Gaines County (Fig. 1B). These results on spatial spread are consistent with reported measles cases of the 2025 measles outbreak in West Texas. Of the 11 West Texas counties that had reported six or more measles cases (twice the CDC’s minimal measles outbreak size [21]) as of June 16th, 2025 [8]), our model showed that nine of them have at least a 0.6 probability of experiencing an outbreak (Fig. 1B & Table S1).

As MMR vaccination is the primary strategy for preventing measles outbreaks, we analyzed the potential impact of increased vaccine uptake on the risk and spatial spread of a Gaines County measles outbreak in Texas. We considered two targeted pre-emptive strategies where vaccine uptake was increased to 90% and 92% in all counties with coverage lower than 90% and 92%, respectively. Under these targeted strategies, the number of counties with outbreak probability greater than or equal to 0.5 was reduced from 14 to 9 (Figs. 2A-B & S3). Each strategy substantially reduced outbreak risk across West Texas, except in Lubbock, Yoakum, Ector, and Midland County, whose risk remained higher than 0.8 (Figs. 2A-B & S2A-B). Both targeted strategies had marginal differences and minimal impact on preventing the geographical spread of a measles outbreak originating in Gaines County (Figs. 2A-B & S2A-B). In addition to the targeted strategies, we considered a uniform pre-emptive strategy where vaccination coverage was increased by 5% in each county (Fig. 2C, S2C & Table S2). This vaccination scenario reduced the number of counties with baseline outbreak risk greater than 0.5 from 14 to 6 counties (Figs. S2& S3). Though all three strategies do not prevent the spread of the measles outbreak across West Texas, our results showed that a statewide increase in MMR coverage is necessary to prevent it from spreading beyond West Texas (Figs. S2 & Table S2). Moreover, a uniform increase in vaccine uptake across counties (strategy 3) was shown to substantially reduce the size of the population at risk of infection (population not effectively immunized) compared to the targeted strategies (Fig. S4).

**Figure 2.**
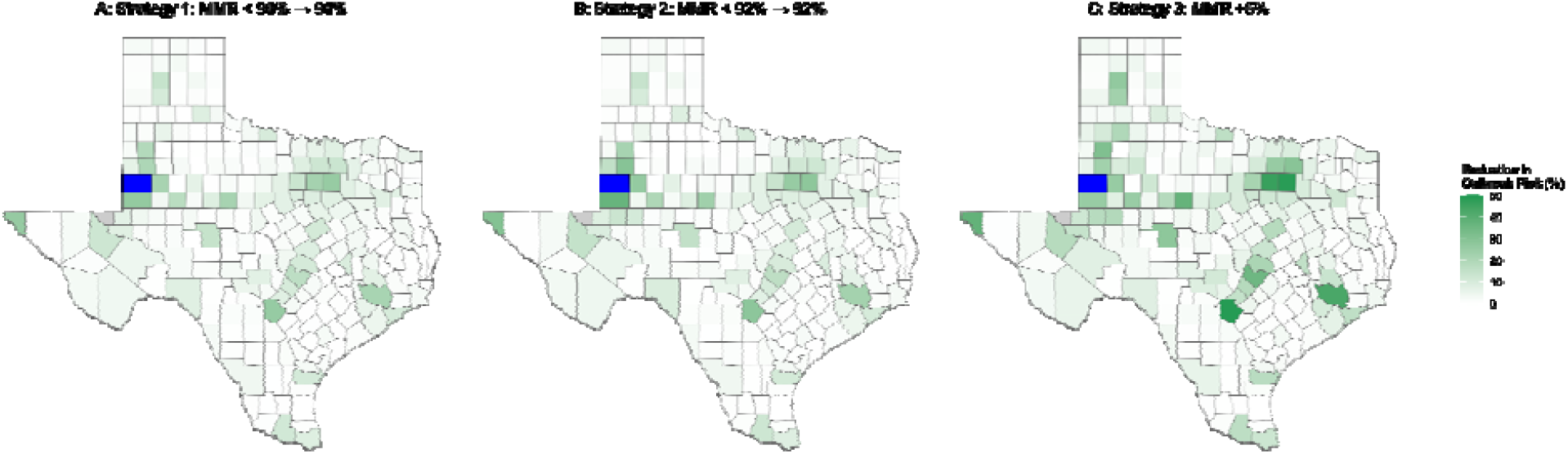
Reduction of the probability that a measles outbreak originating in Gaines County leads to major outbreaks in other Texas counties under three statewide vaccination strategies. Here, the outbreak probability represents the risk of first-generation and second-generation outbreaks. A) Strategy 1: All counties with MMR vaccine coverage below 90% are increased to 90%; B) Strategy 2: All counties with MMR vaccine coverage below 92% are increased to 92%; C) Strategy 3: All counties’ MMR vaccine coverage are increased by 5% (up to 100% vaccine coverage). Gaines County is denoted in blue.

From our baseline MMR vaccine coverage, King County (80.6%), Gaines County (81.7%), Hall County (82.0%), and Throckmorton County (83.4%) were identified as the lowest vaccine coverage counties (Fig. S1 & Table S2). These counties are all located in West Texas. We investigated the spatial spread of a hypothetical 2025 measles outbreak that would have originated in King, Hall, or Throckmorton County. We showed that such outbreaks are unlikely to spread widely to other Texas counties through first- or second-generation outbreaks (Fig. S5). This was likely due to their remote location and limited interaction (number of between-county visits) with other counties. In addition of these West Texas counties, we considered other low vaccine coverage counties such as Limestone (86.4%), Montague (86.8%), and Polk County (89.2%) (Fig. 3). Contrary to King, Hall, and Throckmorton County, these counties are situated in proximity of large metropolitain areas such as Dallas-Fort Worth and Houston. We investigated the spatial spread of outbreaks originating in in these counties, and showed that such outbreaks would have high probabilities of spreading widely and causing local major outbreaks in densely populated areas such as the Houston Metropolitan areas for a Polk County outbreak, the Dallas-Fort Worth Metropolitan area for a Montague County outbreak, and the Dallas-Houston Corridor area for a Limestone outbreak (Fig. 3).

**Figure 3.**
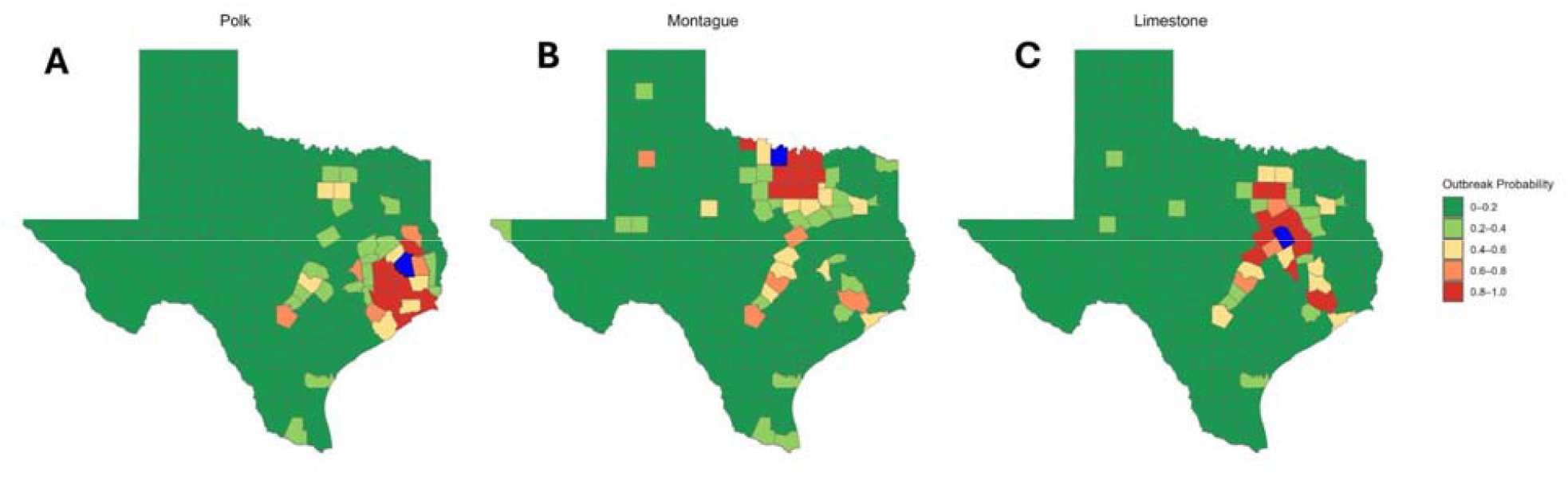
Probability that a measles outbreak originating in Polk, Montague, and Limestone counties leads to major outbreaks in other counties. A) Outbreak originating in Polk County; Polk County is denoted in blue. B) Outbreak originating in Montague County; Montague County is denoted in blue. C) Outbreak originating in Limestone County; Limestone County is denoted in blue. Here, the outbreak probability represents the risk of first-generation and second-generation outbreaks.

## Discussion

We estimated the risk of the spatial spread of a 2025 measles outbreak in Gaines County, Texas. Though such an outbreak can spread beyond state borders, our analysis was limited to the spread across Texas counties. Using county-level MMR vaccination coverage among kindergartners from 2020 to 2024, and empirical data on between-county mobility, we showed that a measles outbreak originating in Gaines County would have at least an 80% probability of directly generating local outbreaks in six neighbouring counties in West Texas (Lubbock, Terry, Yoakum, Andrews, Ector, and Midland County). The risk of the Gaines County outbreak directly generating a local outbreak in other counties was less than 50% in all remaining counties, except Dawson County. These county-level outbreaks resulting from direct contact with infected residents of Gaines County were denoted as first-generation outbreaks. When considering the risk of first-generation and second-generation (outbreaks directly resulting from a first-generation outbreak) outbreaks, we showed that a Gaines County outbreak would have at least a 70% probability of generating local outbreaks in four densely populated counties, including El Paso, Bexar, Tarrant, and Dallas County.

As increased vaccination coverage is the most effective approach for preventing and curtailing the spread of measles, recent modeling studies have investigated the potential impact of vaccination on the risk of measles outbreaks in the US [14,22–24]. We evaluated the impact of increased MMR vaccine coverage on the spread of a measles outbreak originating in Gaines County, Texas. We found that increased routine childhood measles vaccination will substantially reduce the risk and spatial spread of such a measles outbreak across Texas counties. We showed that though targeted strategies that aim to increase vaccine coverage in low coverage counties can substantially reduce the spatial spread of a measles outbreak, a uniform increase of vaccine coverage across counties could be more effective in reducing the number of non-immunized individuals across the state.

MMR vaccination coverage among school-age children is highly heterogeneous across Texas, with several county-level coverages below 90% [16]. Schools in these low-vaccine-coverage counties are regarded as hotspots for measles outbreaks [25,26]. Though the ongoing West Texas measles outbreak, which originated in Gaines County, is one of the largest outbreaks in the measles post-elimination era, our model showed that measles outbreaks in other low vaccination counties, such as Polk, Limestone, and Montague County, could quickly spread to densely populated counties and potentially generate more cases than the current outbreak. Our results highlight a synergistic contribution of vaccination coverage and human mobility to the risk of geographical spread of a measles outbreak in Texas. This result agrees with previous studies showing that human mobility plays a key role in the spread of emerging and re-emerging infectious diseases [27–29].

Our modeling framework estimates the probability of a major outbreak as its main outcome measure. This measure accounts for the potential impact of epidemiological uncertainties in infectious disease transmissions. A county is identified as a high-risk county if it has a high probability of experiencing a major local outbreak. Such a high probability indicates low vaccination coverage and a high risk of case importations due to high contact volume with other high-risk counties. Conversely, a low likelihood of experiencing a major outbreak does not indicate that a county may not report several measles cases during a state-level outbreak. Here, reported measles cases in low-risk counties may simply be imported cases (infection acquired out-of-county) or the result of small outbreaks with limited transmission events [30,31].

Like other modeling studies, our study has several limitations, mostly related to data availability and model assumptions. First, our model used 2019 mobility data on between-county visits, which was informed by the 2019 anonymous mobile phone users’ visits data provided by SafeGraph. These data provide a more accurate description of population mobility than standard mobility models such as the gravity and radiation models [32]. Using prior COVID pandemic mobility data can be regarded as a good approximation of the post-pandemic mobility. Though using 2024 mobility data may have been ideal to inform 2025 mobility, such data are not publicly available. Second, our model uses kindergarten vaccination data to inform the county-level immunization rate, as these data are not publicly available in Texas. Though most measles cases in the US occur among school-age children [26], assuming that the county vaccination rate is equal to its kindergarten coverage ignores potential variation in immunization rates among older children and adults, as well as the within-county spatial clustering of nonvaccinators that may enhance the risk of local outbreaks [33,34]. Future models should refine this assumption to improve the estimation of outbreak risk. Finally, our approach does not account for transient dynamics in outbreak risk, which may be important for forecasting infection cases and hospitalizations. To explicitly account for transient epidemic dynamics, our model can be extended to incorporate within-county/city/school-district disease transmission dynamic models. Calibrating such models would require county/city/district-level time series data on measles cases. However, these time series data are not generally publicly available at the right spatial resolution.

Our results indicate that measles remains a major public health problem in Texas, with the potential risk of causing large-scale statewide outbreaks. This risk is exacerbated by the recent decline in MMR vaccination in the US since the peak of the COVID pandemic [35]. If vaccination rates continue to decline, large-scale measles outbreaks are likely to become frequent in the US in general and in Texas in particular. Our study provides a simple and easy-to-use data-driven modeling framework that can be used to estimate the county-level risk and geographical spread of a measles outbreak and design response strategies for containing or preventing an outbreak. This modeling framework is easily applicable to other US states and can be used at higher spatial resolutions, such as school-district or city-level, conditioned on the availability of vaccination and mobility data.

## Data Availability

All data produced in the present work are contained in the manuscript

**Alt Text Figure 1**: Maps depicting county-level probability of local measles outbreaks originating from Gaines County, Texas.

**Alt Text Figure 2**: Maps depicting the county-level reduction of probability that a measles outbreak originating in Gaines County leads to major outbreaks in other Texas counties under three statewide vaccination scenarios.

**Alt Text Figure 1**: Maps depicting county-level probability that a measles outbreak originating in Polk, Montague, and Limestone counties leads to major outbreaks in other Texas counties.

