## Supplemental materials for "Risk and Spatial Spread of a Measles Outbreak in Texas"

**Supplementary Materials**

**Supplementary figures and Table S1**

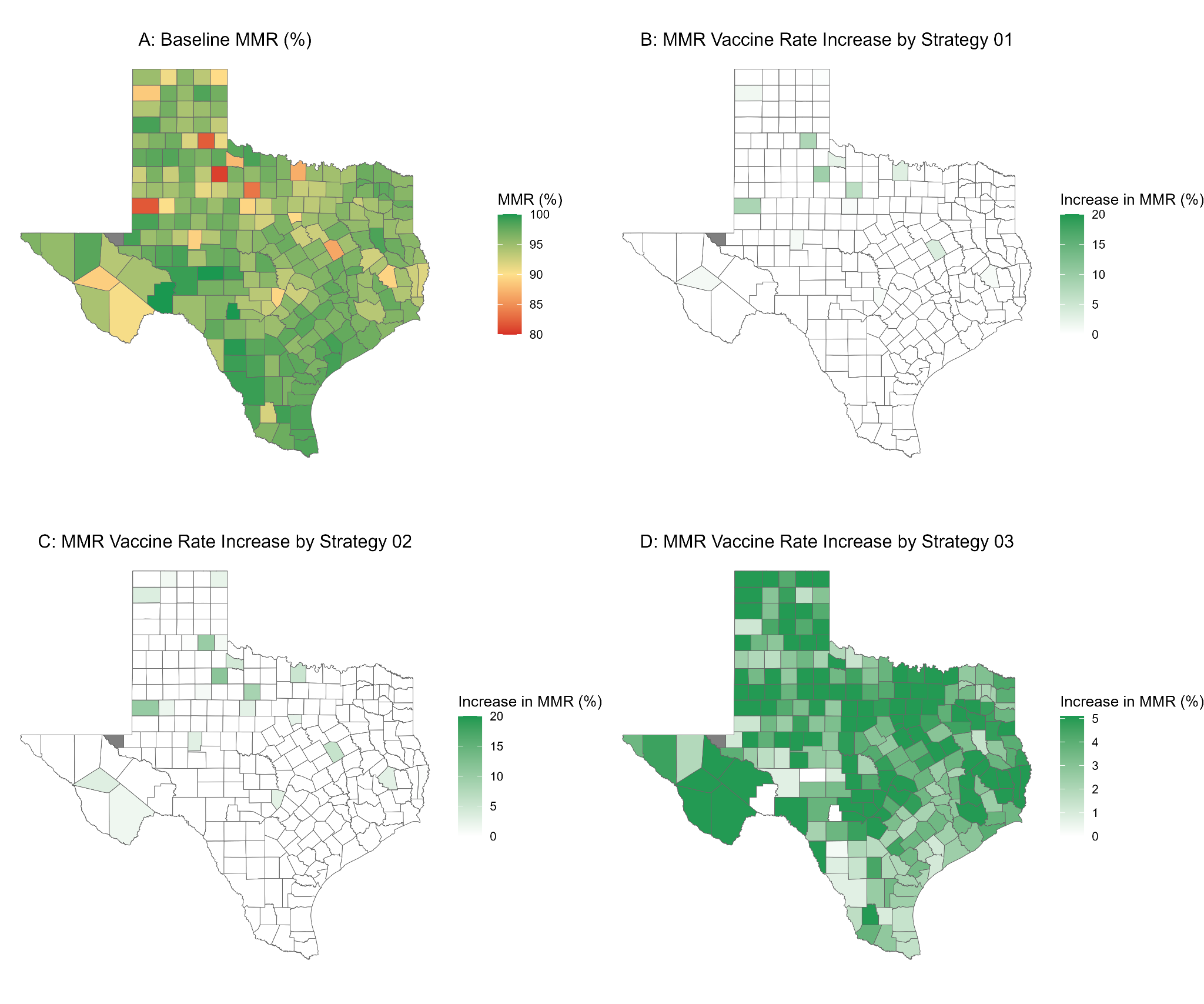

**Figure S1**: County-level vaccination coverage used in our analysis. **A**: The map shows the average county-level vaccination coverage among kindergartners in Texas from 2020 to 2024. **B**: The increase in vaccination coverage under strategy 1, where counties with coverage less than 90% are increased to 90%. **C**: The increase in vaccination coverage under strategy 2, where counties with coverage less than 92% are increased to 92%. **D**: The increase in vaccination coverage under strategy 3, where each county's vaccination coverage would increase by 5% (up to 100% coverage).

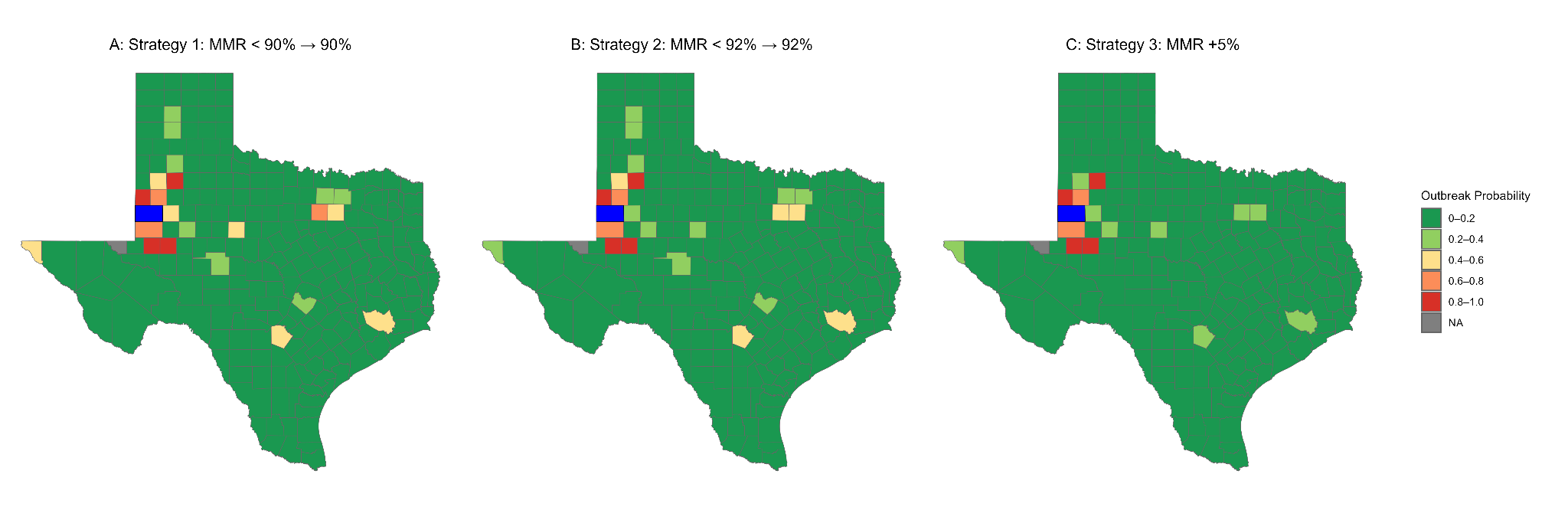

**Figure S2:** Probability that a measles outbreak originating in Gaines County leads to a major outbreak in other Texas counties under three statewide vaccination strategies. Here, the outbreak probability represents the risk of first-generation and second-generation outbreaks. **A**) Strategy 1: All counties with MMR vaccine coverage below 90% are increased to 90%; **B**) Strategy 2: All counties with MMR vaccine coverage below 92% are increased to 92%; **C**) Strategy 3: All counties MMR vaccine coverage are increased by 5% (up to 100% vaccine coverage). Gaines County is denoted in blue.

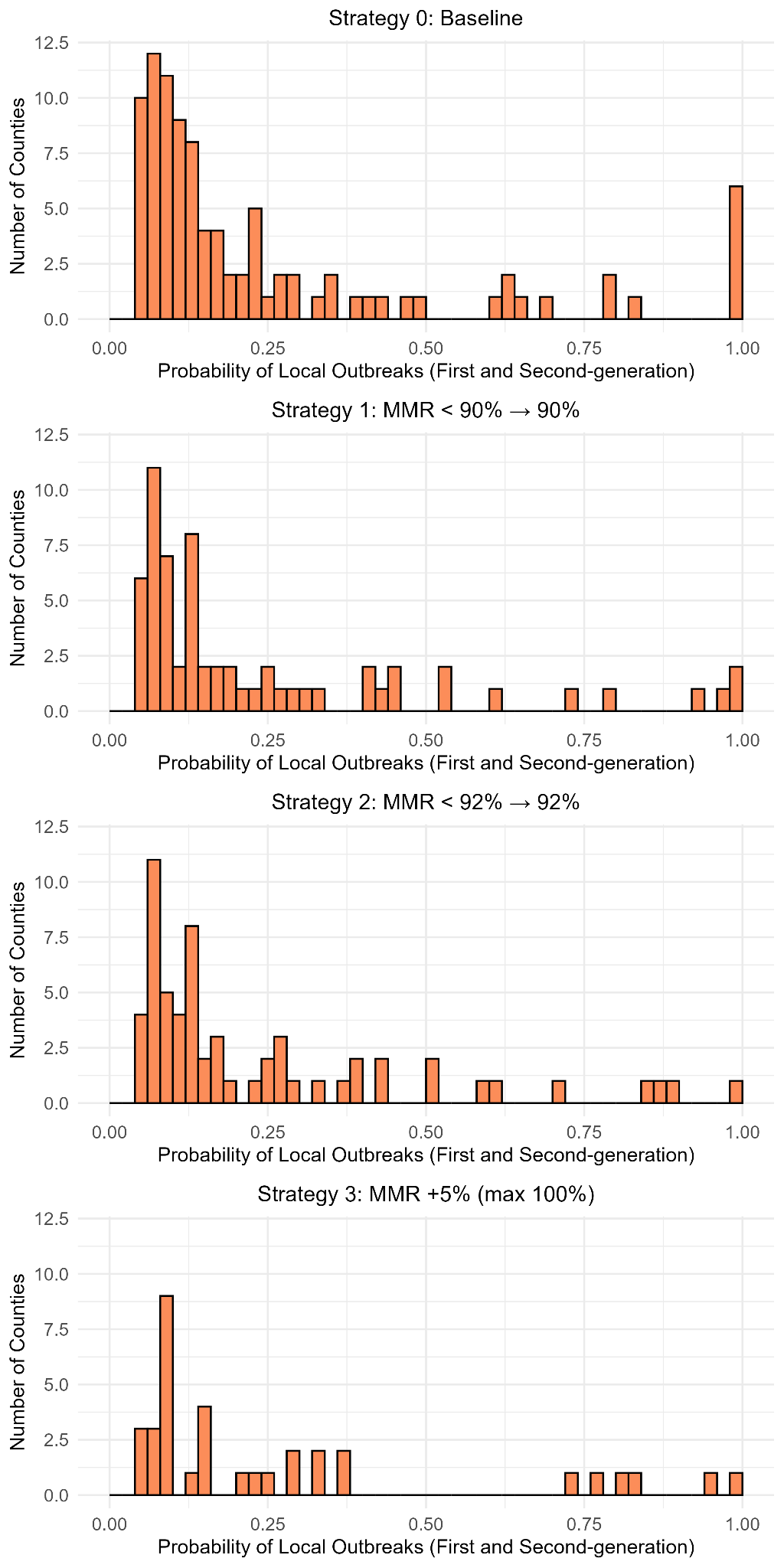

**Figure S3:** Distribution of county-level probability of major outbreaks generated by a measles outbreak originating in Gaines County. Here, probability represents the risk of first-generation and second-generation local outbreaks.

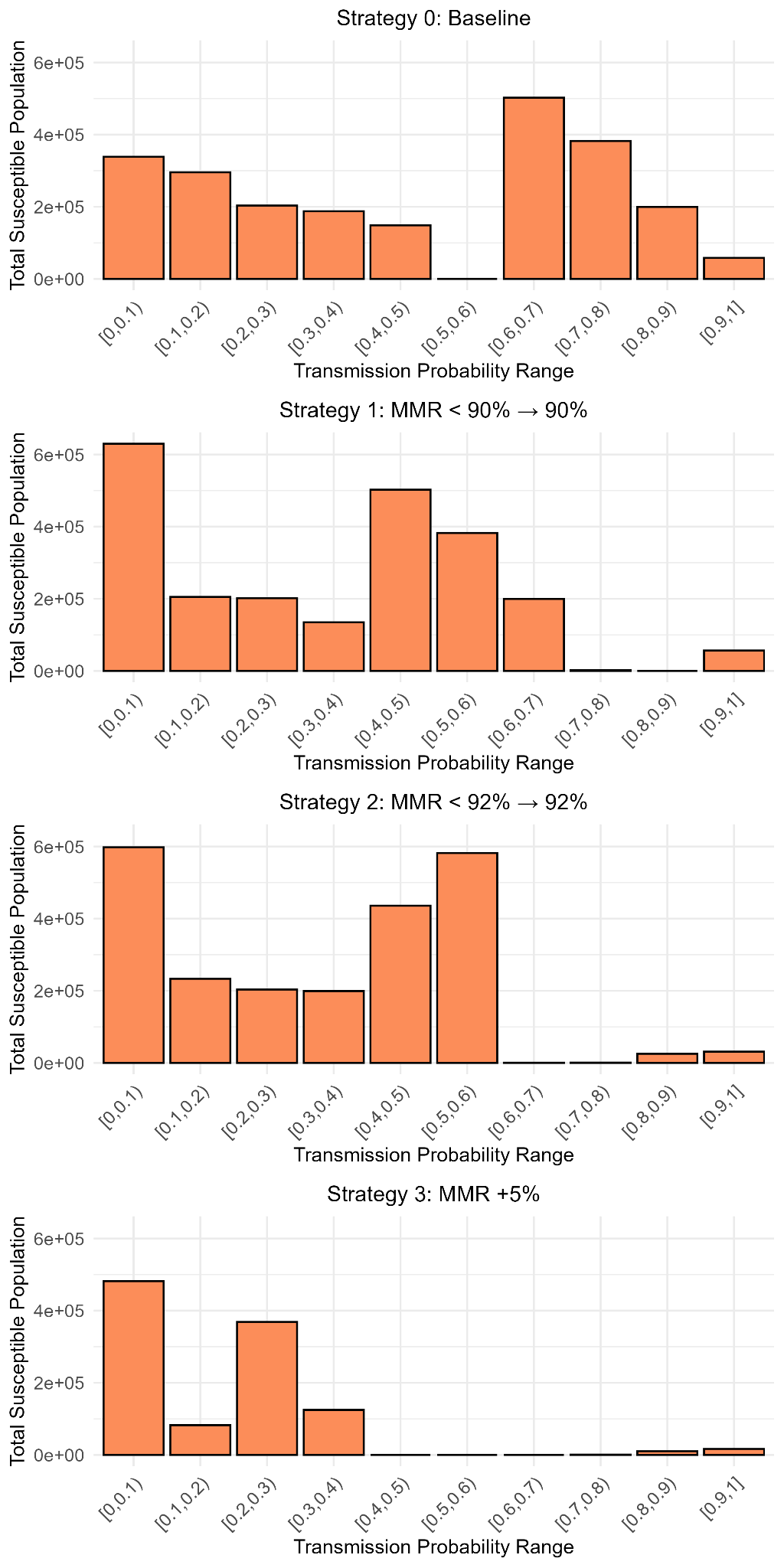

**Figure S4:** Distribution of the susceptible population at risk of experiencing a major outbreak for each vaccination strategy. The susceptible population is defined as the number of individuals unvaccinated or not effectively vaccinated in each county. For a given county *i* with $V_{i}$ MMR vaccination coverage, the proportion of the population not effectively vaccinated is equal to $(1-\epsilon)V_{i}$. Here, probability represents the risk of first-generation and second-generation local outbreaks.

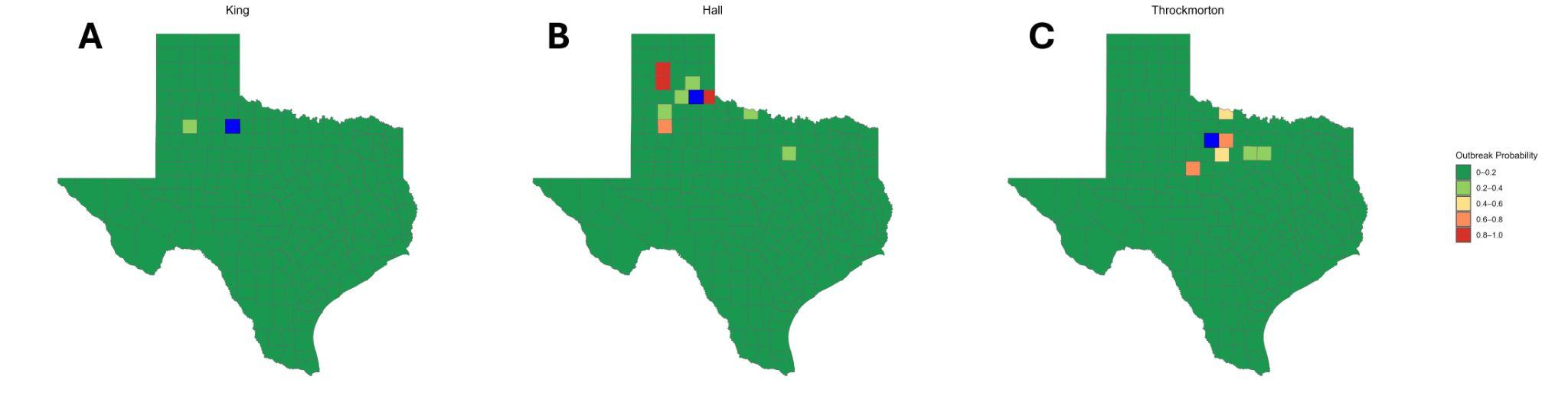

**Figure S5**: Probability that a measles outbreak originating in King, Hall, and Throckmorton County leads to a major outbreak in other counties. **A:** Outbreak originating in King County; King County is denoted in blue. **B**: Outbreak originating in Hall County; Hall County is denoted in blue. **C**: Outbreak originating in Throckmorton County; Throckmorton County is denoted in blue. Here, the outbreak probability represents the risk of first-generation and second-generation outbreaks.

Table S1: Outbreak cases by county in West Texas as of June 16th, 2025

| County name | Confirmed cases |
| --- | --- |
| Dallman | 7 |
| Potter | 1 |
| Carson | 1 |
| Randall | 1 |
| Parmer | 5 |
| Bailey | 2 |
| Lamb | 1 |
| Hale | 5 |
| Cochran | 14 |
| Hockley | 6 |
| Lubbock | 53 |
| Yoakum | 20 |
| Terry | 60 |
| Lynn | 2 |
| Garza | 2 |
| Gaines | 413 |
| Dawson | 26 |
| Andrews | 3 |
| Martin | 3 |
| Ector | 12 |
| Midland | 6 |
| Reeves | 2 |
| Brewster | 1 |
| El Passo | 58 |
